# Magnitude and Associated Factors of Out-of-Pocket Healthcare Expenditure among Outpatients Visiting Public Hospitals in Jigjiga Town, Somali Region, Eastern Ethiopia

**DOI:** 10.64898/2026.03.28.26349597

**Authors:** Mahamed Muhumed Ahmed, Desalegn Demeke Shitaye, Abera Cheru, Adisu Birhanu Weldesenbet, Belay Negash

## Abstract

**Background:** Out-of-pocket healthcare expenditure (OOPHE) remains a major challenge to accessing adequate medical service, often discouraging individuals from seeking necessary medical services. The extent of OOPHE in Jigjiga city is unknown. This study aimed to assess the magnitude and associated factors of OOPHE among outpatients visiting public hospitals in Jigjiga city, Somali region, Eastern Ethiopia.

**Methods:** A hospital-based cross-sectional study was conducted among 406 outpatients selected through systematic random sampling from three public hospitals in Jigjiga city. Data were collected through interviews-administered questionnaires and analysed by SPSS version 25.0. Binary and multivariable logistic regression analyses were performed to identify factors associated with OOPHE among outpatients (p < 0.05).

**Results:** Overall, 89.5% of respondents incurred out-of-pocket healthcare payments at the point of service delivery. The mean OOPHE per outpatient was 485.6 ± 349 birr ($3.12 ± $2.24). Female [AOR = 3.38, 95% CI (1.54-7.42)], unmarried [AOR = 5.32, 95% CI (1.77-16.03)], and traveled ≥5 km [AOR = 7.07, 95% CI (1.46-34.29)] and higher educational attainment (college and above) [AOR = 7.07, 95% CI (1.55-32.28)] were independently associated with higher odds of incurred OOPHE.

**Conclusion:** The magnitude of out-of-pocket healthcare payments among outpatients was high. Sex, marital status, educational level, and distance to reach a public health facility were significant predictors of OOPHE. Policy action to reduce OOPHE in this setting should include strengthening and expanding the Community-Based Health Insurance scheme and promoting prepayment mechanisms, such as Social Health Insurance, for formal sector employees, specifically for government employees.

## Introduction

Out-of-pocket health expenditure (OOPHE) is defined as household spending incurred when using a service to obtain any type of health care for motivated, preventive, curative, rehabilitative, palliative, or long-term reasons (1). High OOPHE can force individuals to discontinue treatment or even avoid seeking health services, among others, due to being unable to afford these costs (2). Globally, each year around 150 million people from 44 million households face the economic burden of OOPHE, and nearly more than 100 million people from 25 million households are placed into poverty as a result of OOPHE (3).

According to the World Health Organization, catastrophic out-of-pocket health expenditure occurs when direct out-of-pocket payments exceed either 10% of total household income or 40% of household income minus subsistence needs (4). The World Health Organization initiated Universal Health Coverage, aiming to ensure that every person has access to health care without suffering financial hardship (5), through shifting out-of-pocket payment to prepayment system like Community-Based Health Insurance (CBHI) (6). Despite these efforts, millions of households in low- and middle-income countries are still at risk of financial hardship due to out-of-pocket healthcare costs (7).

In most low- and middle-income countries, OOPHE accounts for 20% to 60% of national health expenditure while in most developed economies, this amount accounts for only 15% to 25% of OOPHE (8). In Africa, out-of-pocket payments account for more than 25% of total healthcare spending in 31 countries, 50% in 11 countries, and 70% in 3 countries (4). In Ethiopia, out-of-pocket payments account for about 31% of total health expenditure, which is greater than 20% of the global recommendation target and poses a significant obstacle to equitable access to healthcare (9). Many households borrow money, sell assets, or shift resources from other requirements to access healthcare. In fact, high OOPHE in the absence of risk-pooling mechanisms and a high degree of poverty can result in deep and catastrophic financial shocks to vulnerable households (10).

In Ethiopia, OOPHE represents a substantial portion of total healthcare spending that creates a significant financial barrier to accessing adequate healthcare and may deter individuals from seeking necessary medical services (11). The Somali region, one of Ethiopia’s four Developing Regional States, is predominantly inhabited by pastoralists who experience developmental inequities and poorer health outcomes compared to the national average (12). In Jigjiga town, many households are complaining about the rapid growth of healthcare costs incurred at the point of service delivery. This payment system causes a huge financial burden that leads to impoverishment. However, there is no any relevant study conducted in the region at all, so the purpose of this study is designed to assess the magnitude and associated factors of OOP medical expenditures among outpatients visiting public hospitals in Jigjiga town, Somali region, Eastern Ethiopia, and improve the general structure of financial risk pooling schemes as well as provide updated baseline information for health planners on how to intervene.

## Methods and Materials

### Study area and period

A hospital-based cross-sectional study was conducted at Sheik Hassan Yabare Comprehensive Specialized Hospital, Karamara General Hospital (KGH), and Jigjiga Primary Hospital (JPH). These facilities are located in Jigjiga city, the capital city of the Somali Regional State in Eastern Ethiopia, which is located 640 km away from Addis Ababa, the capital city of Ethiopia. Jigjiga city has one comprehensive specialized hospital, one general hospital, one primary hospital, three health centers, and many private clinics that provide healthcare services to the community (13, 14). The study was conducted from November 15 to 30, 2023.

### Population and Eligibility

The source population was all clients/patients attending outpatient departments in public hospitals of Jigjiga city, Somali region whereas, all selected individuals who received outpatient services from the public hospitals were the study population. All outpatient clients present at the time of data collection were included in the study and patients with serious medical illnesses, that prevent them from responding to the questionnaire and whose residential areas were out of the region were excluded from the study.

### Sample Size Determination

The sample size was calculated by using EPI info version 7, considering factors that are significantly associated with outcome variables obtained from previous studies. The following assumptions were applied: a two-sided confidence level of 95%, a power of 80%, a ratio of 1:1, and an anticipated 10% non-response rate. Finally, the largest sample size, which is 406, was obtained by using age as a significant factor from the previous study (6).

### Sampling Procedure and Technique

A systematic random sampling technique was used to select the study population. All three public hospitals in Jigjiga town were included in the study. Initially, a comprehensive list of all outpatient departments (clinics) was created and the average number of patients who visited each outpatient department for the same period of our data collection from last year’s hospitals’ reports was assessed. The sampling interval (K) was determined by dividing the total number of outpatients attending public hospitals by the final sample size, and the calculated sample size was proportionally allocated to each hospital. Using this approach, a total of 406 outpatients were systematically selected with an interval of 16. Of these, 122 outpatients were recruited from Karamara General Hospital, 184 were recruited from Sultan Yabare Referral, and 100 were recruited from Jigjiga Primary Hospital **(Figure 1)**.

**Figure 1:**
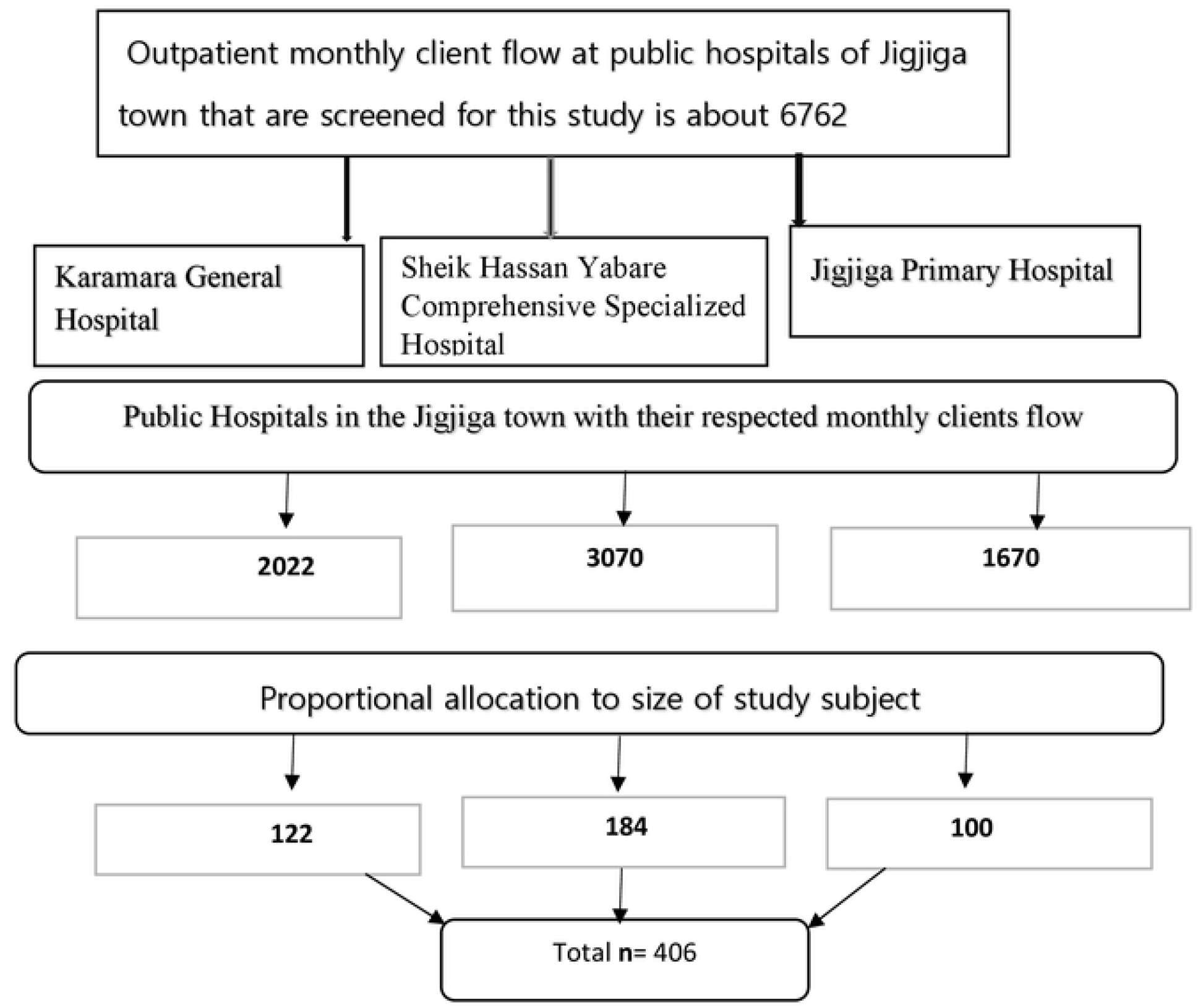
Sampling procedure on magnitude of *OOPHE* and associated faactors among outpatients in public hospitals Jigjiga Town Somali Region, Eastern Ethiopia. Note: OOPHE Out of pocket health expenditure, CBHI-Community based health insurance

### Data Collection Instrument

The data collection tool was developed by review of previous literatures (6, 11, 15). The contents of the tool include socio-demographic factors and economic characteristics, Patients’/Households’ Health insurance status, patients’ household food consumption and OOPHE made by Patients/Patients’ households’. Data collection was performed by data collector through interview-administered questionnaires. The questionnaire was developed in English, translated into the local language Somali, and back-translated to English by language experts to ensure consistency. The validity and reliability of the Somali version of the questionnaire were confirmed.

### Data Collection Method and Procedure

Data were collected by four trained public health professionals under the close supervision of the principal investigator. Prior to data collection, the data collectors received one day of training on research ethics, data collection procedures, and the administration of the questionnaire. Eligible clients attending outpatient rooms in the selected hospitals during the survey period were approached. The study objectives were explained to them, and an interview was conducted only with those who gave their consent. Outpatient clients were the primary responders in the survey; however, when a client was unable to provide the necessary information, particularly regarding cost, her partner/family members stepped in to help.

### Study Variables

The dependent variable was out-of-pocket healthcare expenditure among outpatients, while the independent variable included socio-demographic factors such as age, sex, educational status, marital status, and family size, along with economic status, distance to the hospital, type of illness, frequency of hospital visits, insurance status, type of insurance, and average payment.

### Operational Definitions

**Out-of-pocket healthcare expenditures** are determined as the proportion of respondents who faced out-of-pocket payment for consultation, medication, and an investigation among participants who received the outpatient services during the data collection period (6).

### Direct non-medical cost

Non-medical care expenditure includes total transportation costs from home to hospital and food consumed for both client and their caregiver (companion).

### Data Quality Control

To ensure high-quality data, one day of training was given to all data collectors and supervisors about all aspects of the data collection tools, questioning techniques, and ethical issues by the principal investigator. A pre-test was conducted on 5% of the sample size at Kebribeyah Primary Hospital. During the pretest, the questionnaire was checked for its clarity, simplicity, understandability, completeness, consistency, and coherency. Daily supervision was done by the supervisor and principal investigator.

### Data Analysis

The collected data were rechecked, cleaned, coded, and entered into EPI data version 4.7 and exported to Statistical Package for Social Science (SPSS) version 25 for analysis. Descriptive statistics were done and presented in the form of texts, tables, and figures for categorical variables, whereas continuous variables were summarized by mean with standard deviation (SD) or median. Binary logistic regression analysis was performed, and a crude odds ratio (COR) was computed at a 95% C.I. Thus, variables with a p-value less than 0.25 were considered as potential candidates for the final multivariable logistic regression analysis. Necessary assumptions of logistic regression were checked and a multivariable logistic regression analysis was subsequently run to identify factors associated with OOPHE among outpatients. Statistical significance was declared at p-value of less than 0.05.

### Ethical Consideration and Consent Participants

Ethical approval for the study was obtained from the ethical review committee of Haramaya University, College of Health and Medical Sciences (institutional health research ethics review committee Ref. No. 206/2023). Formal letters for cooperation were obtained from Haramaya University, College of Health and Medical Science, and submitted to Jigjiga city administration and public hospitals in Jigjiga town. Informed, volunteer, written, and signed consent was obtained from the study participants after they were informed in detail about the objective, purpose, benefits, and risks of the study. The study was maintained confidentially at all levels.

## Results

### Socio-demographic and economic characteristics of study participants

A total of 400 outpatients seeking health services participated in the study, with a response rate of 98.5%. The mean age of participants was 36.9 years with an SD of 13.8. Among the total participants, 214 (53.5%) were male, 309 (77.2%) were urban dwellers, and 85 (21.2%) had attained college and above. Less than half of the participants, 187 (46.8%) had a monthly income between 5000 and 10000 ETB ($32.13 and $64.26), with a mean monthly income of 12111.88 ± 8915.94 ETB ($73.83 ± $57.3) **(Table 1)**.

**Table 1:** Socio-demographic and economic characteristics of study participants attending a public hospital in Jigjiga Town, Somali Region, Eastern Ethiopia.

| Variables | Categories | Number (%) |
| --- | --- | --- |
| Residence | Rural | 91 (22.8) |
|  | Urban | 309 (77.2) |
| Sex | Male | 214 (53.5) |
|  | Female | 186 (46.5) |
| Age | <31 | 165 (41.3) |
|  | 31– 49 | 163 (40.7) |
|  | ≥ 50 | 72 (18) |
| Marital status | Unmarried | 120 (30.0) |
|  | Married | 280 (70.0) |
| Educational status | cannot read and write | 160 (40) |
|  | Grade 1-8 | 76 (19) |
|  | Grade 9-12 | 79 (19.8) |
|  | College and above | 85 (21.2) |
| Monthly income of HH (ETB) | <5000 | 46 (11.5) |
|  | 5000-10000 | 187 (46.8) |
|  | ≥10000 | 167 (41.7) |
| Family size | <5 | 106 (26.5) |
|  | ≥5 | 294 (73.5) |

Note: ETB-Ethiopian birr

### Magnitude of out-of-pocket health expenditures among outpatients

Among the 400 participants, 358 (89.5%) incurred out-of-pocket payments to access medical services at the point of service delivery, whereas only 29 (7.2%) were covered by a health insurance scheme **(Figure 2)**. The mean OOPHE was 485.6 ± 349 birr ($3.12 ± $2.24) per participant. The mean expenditure for direct non-medical cost averaged 140.8±185.31 birr per participant **(Table 2)**.

**Table 2:** The mean of OOPHE for outpatient services in public hospitals of Jigjiga city, Somali Region, Eastern Ethiopia, (in ETB).

| Type of cost | Mean (ETB) | Median (ETB) |
| --- | --- | --- |
| Investigation cost | 218.2(SD 205.63) | 150 |
| Consultation cost | 14.1 (SD 9.87) | 10 |
| Medication cost | 253.3 (SD 209.46) | 200 |
| Transportation cost | 83.7 (SD 115.77) | 50 |
| Food cost | 56.8 (SD 69.54) | 40 |
| Total OOP healthcare cost | 485.6 (SD 349) | 440 |

Note: ETB-Ethiopian birr

**Figure 2:**
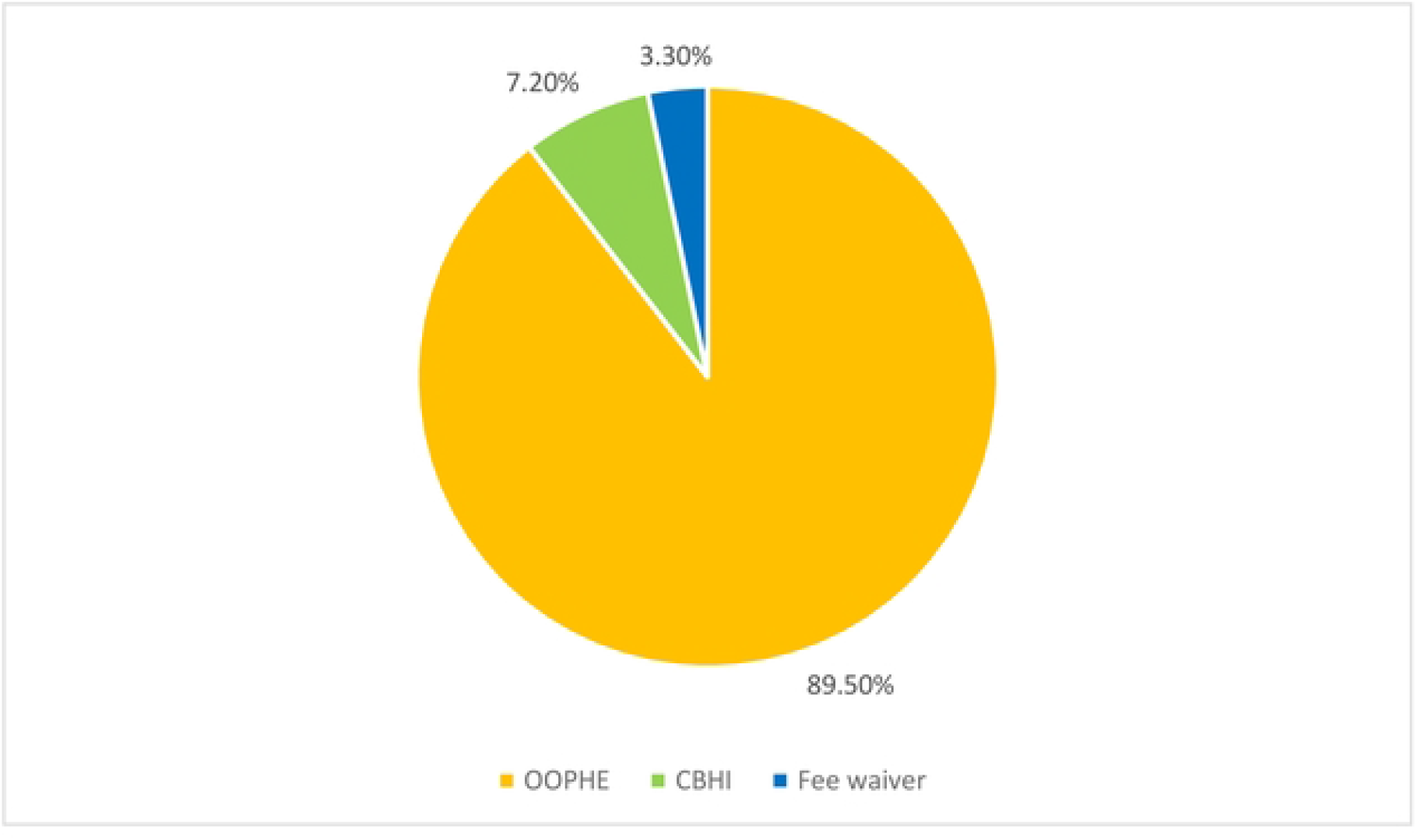
Magnitude of OOPHE among participants visiting outpatients in public hospitals of Jigjiga City Somali Region, Eastern Ethiopia Note: OOPHE-Out-of-pocket health expenditure, CBHI-Community based health insurance

### Clinical and related factors

Two hundred and forty-one (60.2%) of the participants received diagnoses for acute illnesses, while 106 (26.5%) received diagnoses for chronic illnesses. In terms of access to healthcare facilities, around half of the participants 203 (50.7%), traveled less than five kilometers to reach public health facilities **(Table 3)**. Regarding healthcare financing, more than half of participants 221 (55.3%), reported family support as their primary source of funds for OOPHE **(Figure 3)**.

**Table 3:**
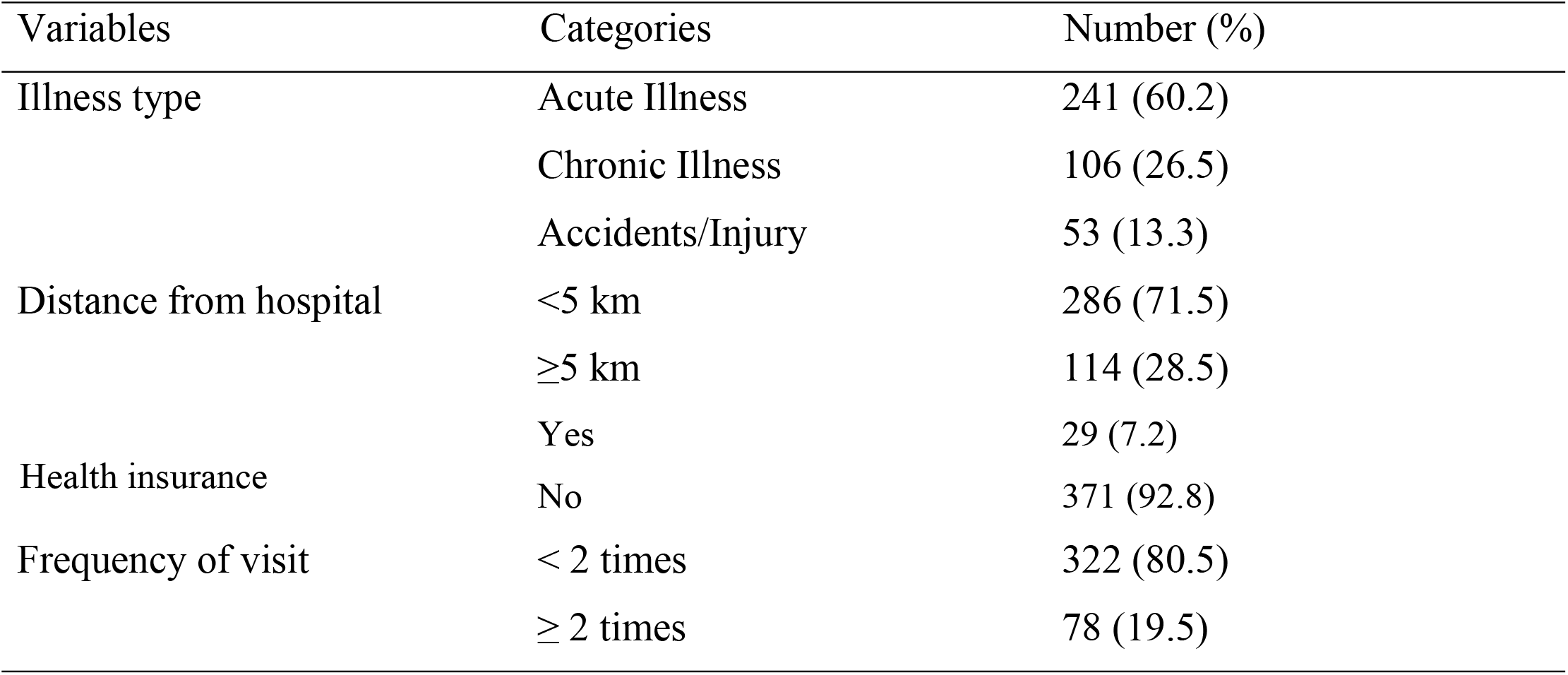
Clinical and related factors of study participants attending a public hospital in Jigjiga Town, Somali Region, Eastern Ethiopia.

**Figure 3:**
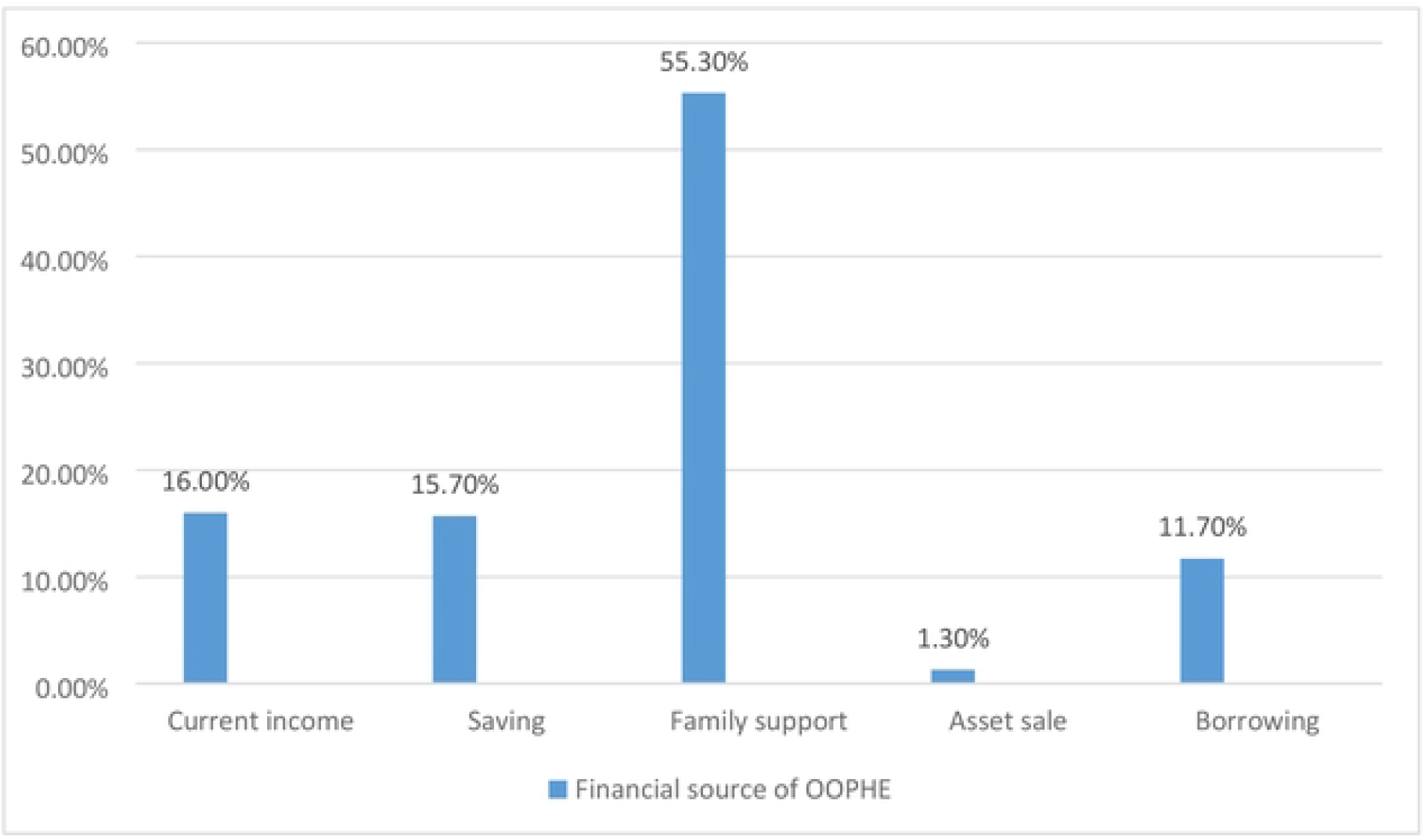
Financial source of OOPHE among participants visiting outpatients in public hospitals of Jigjiga City, Somali Region, Eastern Ethiopia

### Factors associated with out-of-pocket health expenditures

In the bivariate logistic regression analysis, variables with a p-value less than 0.25 (sex, residence, marital status, educational status, family size, frequency of hospital visits per month, and distance from health facility) were included in the multivariate logistic regression analysis to identify significant predictor variables of OOPHE. The multivariate logistic regression analysis indicated that four variables were significantly associated with OOPHE. Study participants who were female were 3.4 times more likely to pay OOPHE than male [AOR = 3.38, 95% CI (1.54-7.42)]. Regarding marital status, unmarried participants were 5 times more likely to pay OOPHE than married participants [AOR = 5.32, 95% CI (1.77-16.03)]. With respect to educational status, those who had completed college and above were 7 times more likely to pay OOPHE compared to participants who could not read and write [AOR = 7.07, 95% CI (1.55-32.28)]. Furthermore, participants who traveled ≥5 km to reach a public health facility had 7 times higher odd of incurring OOPHE compared with those who traveled <5 km [AOR = 7.07, 95% CI (1.46-34.29)] **(Table 4)**.

**Table 4:** Binary and multivariable logistic regression analysis of factors associated with OOPHE among outpatients in public hospitals of Jigjiga city, Somali Region, Eastern Ethiopia.

| Variable | Category | OOPHE |  | COR (95%CI) | AOR(95%CI) |
| --- | --- | --- | --- | --- | --- |
|  |  | Yes (%) | No (%) |  |  |
| Sex | Male | 183(85.5%) | 31(14.5%) | 1 |  |
|  | Female | 175(94%) | 11(6%) | 2.69 (1.31-5.53) | **3.38(1.54-7.42) \*\*** |
| Residence | Rural | 88 (96.7%) | 3 (3.3%) | 4.24 (1.28-14.05) | 1.88(0.48-7.29) |
|  | Urban | 270 (87.3%) | 39(12.7%) | 1 |  |
| Marital Status | Unmarried | 116(96.6%) | 4(3.4%) | 4.55 (1.59-13.06) | **5.32(1.77-16.03) \*\*** |
|  | Married | 242(86.4%) | 38(13.6%) | 1 |  |
| Educational status | Can't read & write | 139(86.8%) | 21(13.2%) | 1 |  |
|  | Grade 1-8 | 65(85.5%) | 11(14.5%) | 0.9(0.41-1.96) | 1.18(0.50-2.80) |
|  | Grade 9-12 | 71(89.8%) | 8(10.2%) | 1.3(0.56-3.18) | 1.62(0.64-4.12) |
|  | College & above | 83(97.6%) | 2(2.4%) | 6.3(1.43-27.42) | **7.07(1.55-32.28) \*** |
| Family size | <5 | 90(85%) | 16(15%) | 0.55(0.28-1.07) | 0.51(0.24-1.08) |
|  | ≥5 | 266(91.1%) | 26(8.9%) | 1 |  |
| Frequency of visit per month | < 2 | 282(87.6%) | 40(12.4%) | 0.18(0.04-0.78) | 0.24(0.05-1.06) |
|  | ≥ 2 | 76(97.4%) | 2(2.6%) | 1 |  |
| Distance from health facility | <5 km | 246(86%) | 40(14%) | 1 |  |
|  | ≥ 5 KM | 112(98.2) | 2(1.8%) | 9.12(2.12-38.34) | **7.07(1.46-34.29) \*** |

Note: \*P < 0.05; \*\*P < 0.01, and \*\*\*P < 0.001; AOR = adjusted odds ratio; COR = Crude odds ratio; OOPHE = Out-of-Pocket health expenditure

## Discussion

The main objective of this study was to determine the magnitude and factors associated with out-of-pocket health expenditure (OOPHE) among patients attending public hospitals in Jigjiga City, Somali Region, Eastern Ethiopia. The finding revealed that 89.5% of respondents incurred out-of-pocket payments at the point of service delivery. This finding is consistent with a study conducted in East Shon Zone Public Hospital Ethiopia, which reported a value of 87.8% (6). However, it is lower compared to the study conducted in Nigeria (93.2%) (16), but higher compared to the studies conducted in Colombia (73.9%) (17), India (60.3%) (18), and Cotê d’Ivoire (34.13%) (19). The variation might be due to differences in socio-economic characteristics, study period, setting, health financing and insurance coverage, and geographical variation. The mean OOPHE was $3.12 ± $2.24 per participant. This finding is consistent with the study conducted in Addis Ababa, Ethiopia ($3.92 ± $6.7) (15), but lower compared to the studies conducted among hypertension patients in Shewa Zone, Ethiopia ($22.3) (20), Debre-Tabor, Ethiopia ($113.40 ± $10.18) (21), and Bangladesh ($27.66) (10). Variations in study population characteristics, study period, disease profiles, service utilization patterns, and health financing mechanisms may account for this discrepancy.

In this study, participants who were female were 3.4 times more likely to pay OOPHE than male. This finding is supported by the studies conducted in Debre-Tabor, Ethiopia (21), and USA (22). This is due to the fact that, women typically have higher health-care utilization, owing in part to reproductive and maternal health needs that increase contact with health services and opportunities for OOP payments. Regarding marital status, unmarried participants were 5 times more likely to pay OOPHE than married participants. This finding is consistent with the studies conducted in Gondar (2), Addis Ababa (15) Ethiopia, and Bangladesh (10). Community-Based Health Insurance (CBHI) enrollment in Ethiopia is typically organized at the household level, potentially providing married participants with greater financial protection compared to unmarried participants, who may be less likely to enroll independently. Furthermore, limited financial risk-sharing, reduced social support, and lower insurance coverage among unmarried individuals expose them to OOPHE.

This study found that participants who had completed college and above were 7 times more likely to pay OOPHE compared to participants who could not read and write. This finding is supported by the study conducted in East Shoa Zone, Ethiopia (6), Nigeria (23), and Malaysia (24). A possible explanation for this finding is that individuals with higher education are more likely to be employed in the formal sector, including government positions, and therefore may not be enrolled in CBHI. In the absence of a comprehensive social health insurance (SHI) scheme, these formal-sector workers can remain exposed to substantial OOPHE. Additionally, greater health-seeking behavior and health literacy among the educated may increase service use and acceptance of recommended diagnostics and treatments that further raise OOPHE.

This study revealed that participants who traveled ≥5 km to reach a public health facility had 7 times higher odd of incurring OOPHE compared with those who traveled <5 km. This finding is supported by the study conducted in Debre-Tabor, Ethiopia (21). These finding highlights the combined effect of limited insurance coverage and geographical barriers in exacerbating the financial burden of healthcare utilization.

## Limitations

The facility-based design may have excluded individuals who did not seek care at public hospitals (including non-users and private-sector users), limiting the generalizability of findings to the broader population. As the data was collected in a limited period of time, possible seasonal variations, which may affect the level of cost incurred by outpatients, may not be fully captured by the current findings.

## Conclusion

The study concludes that the burden of out-of-pocket healthcare expenditure among patients attending public hospitals in Jigjiga City, Somali Region, Eastern Ethiopia was high. The study identified that sex, marital status, educational level, and distance to reach a public health facility were significantly associated with OOPHE. To mitigate these financial barriers policymakers should scale-up and strengthening of CBHI scheme to increase enrollment and improve financial protection particularly for vulnerable groups. Additionally, expansion of prepayment mechanisms, such as social health insurance, to reduce the financial burden among formal sector employees, specifically for government employees.

## Data Availability

All relevant data are included within the manuscript and its figures

## Acronyms

AOR: Adjusted odds ratio
CBHI: Community-Based Health Insurance
OOP: Out-of-Pocket Payments
OOPHE: Out-of-Pocket health expenditure

## Declarations

### Ethical approval and Informed consent

This study was conducted in accordance with the principles of the Helsinki Declaration, and ethical clearance was obtained from the Institutional Health Research Ethics Review Committee (IHRERC/206/2023) of the College of Health and Medical Sciences, Haramaya University. A written support letter from Haramaya University was sent to Jigjiga city administration and public hospitals in Jigjiga town. Informed, voluntary, written and signed consents was obtained from all participants.

### Data availability

The datasets used and/or analyzed during the current study are available from the corresponding author on reasonable request.

### Funding Statement

The authors did not receive any funding for this project or the publication of this article.

### Competing interests

The authors declare that there are no competing of interest.

### Author contributions

MMA was involved from inception to the design, acquisition of data, analysis, and interpretation, and drafting and editing of the manuscript. DDS, AC, ABW and BN were the co-authors who participated in the review of the article, tool evaluation, interpretation, and critical review of the draft manuscript. All authors have read and approved the final manuscript.

## Acknowledgment

We sincerely acknowledge the administrative support from Haramaya University, College of Health and Medical Sciences, School of Public Health. We also extend our deepest gratitude to public hospitals in Jigjiga City, Somali region health bureau for granting permission to conduct this study and to all participants for their cooperation and active involvement.

## Notes

### Competing Interest Statement

The authors have declared no competing interest.

### Funding Statement

The author(s) received no specific funding for this work.

### Author Declarations

ethical clearance was obtained from the Institutional Health Research Ethics Review Committee (IHRERC/206/2023) of the College of Health and Medical Sciences, Haramaya University.

